# Cortical hemodynamic biomarkers forecast rTMS responsiveness in post-stroke upper limb motor recovery

**DOI:** 10.1101/2025.05.12.25327481

**Authors:** Xiangbo Wu, Tao Han, Lina Liang, Qun Shi, Mulan Xu, Xiaodong Lin, Baijie Xue, Ming Gao, Xin Zhang, Jiqiang Xie, Xiaolong Sun, Hua Yuan

**Author notes:** **Corresponding to:** Hua Yuan, MD, Department of Rehabilitation Medicine, Xijing Hospital, Air Force Medical University (Fourth Military Medical University), Xi’an, Shaanxi, 710032, P. R. China,; or Xiaolong Sun, MD, Department of Rehabilitation Medicine, Xijing Hospital, Air Force Medical University (Fourth Military Medical University), Xi’an, Shaanxi, 710032, P. R. China. X.Wu, T.Han and L.Liang contributed equally.

## Abstract

**BACKGROUND:** Stroke commonly causes upper limb impairment, significantly reducing quality of life. While low-frequency repetitive transcranial magnetic stimulation (LF-rTMS) over contralesional primary motor cortex (M1) improves arm function, its effectiveness varies, particularly in severe cases.

**METHODS:** Stroke patients with upper limb paralysis were recruited for 14-day LF-rTMS. Baseline data, including demographics and stroke details, were recorded. Upper limb motor function was assessed using the upper extremity Fugl-Meyer assessment (UEFM) and the Wolf motor function test. Functional near-infrared spectroscopy (fNIRS) data were collected during fist clenching, and the laterality index (LI) was calculated to assess the asymmetry of hemispheric activation in the brain. A model was developed and validated both internally and externally to predict the responsiveness of LF-rTMS.

**RESULTS:** This prospective multicenter study included 111 patients with stroke (training cohort, 62; internal/external validation cohorts, 25/24). After 14-day of LF-rTMS, upper limb motor function improved significantly (*P*<0.001), with a response rate of 61.3%. The least absolute shrinkage and selection operator (LASSO) and multivariate logistic regression analyses revealed that the pre-intervention UEFM score (*P*=0.015, odds ratio [OR]=1.048) and pre-intervention LI for M1 for the affected hand (AFF-LI-M1-pre) (*P*=0.001, OR=0.117) were independent predictive factors for the responsiveness of LF-rTMS. The predictive nomogram integrating pre-intervention UEFM score and AFF-LI-M1-pre demonstrated robust discriminative accuracy across cohorts, with AUC values of 0.861 (95% CI 0.770–0.952) in training, 0.853 (95% CI 0.703-1.000) in internal validation, and 0.828 (95% CI 0.645-1.000) in external validation cohorts. Model calibration showed excellent agreement between predicted and observed outcomes (Hosmer-Lemeshow *P*=0.801). Decision curve analysis (DCA) confirmed clinical utility within broad probability thresholds (0.02-0.94), yielding net benefit advantages of 7%-38% over default strategies.

**CONCLUSIONS:** LF-rTMS improved upper limb motor recovery in patients with stroke, with the baseline UEFM score and fNIRS-derived AFF-LI-M1-pre serving as robust predictors. Validated nomogram personalizes therapy, highlighting fNIRS’s role in optimizing treatment. These findings support precision rehabilitation approaches for improving outcomes in resource-limited settings.

**GRAPHIC ABSTRACT:** A graphic abstract is available for this article.

---

Stroke remains a leading global health concern due to its high incidence and mortality rates. Although advancements in diagnosis and treatment have reduced stroke-related mortality, the disability rate remains alarmingly high. Research indicates that approximately 81% of stroke patients suffer from motor impairments, with >60% experiencing upper limb dysfunction, especially concerning hand dexterity and coordination.^1^ These impairments significantly limit patients’ ability to perform self-care tasks, thereby reducing their independence and overall quality of life, as well as imposing substantial burdens on healthcare systems.^2^

Recently, repetitive transcranial magnetic stimulation (rTMS), a noninvasive brain stimulation technique, has demonstrated effectiveness in improving upper limb motor function in patients with stroke.^3^ rTMS modulates cortical excitability in targeted brain regions. High-frequency rTMS (HF-rTMS), which is ≥5Hz, improves cortical excitability, whereas low-frequency rTMS (LF-rTMS), which is ≤1Hz, reduces it.^4^ rTMS facilitates neuroplasticity by promoting neural connections in residual pathways, thereby supporting functional recovery. The classic rTMS stimulation protocol on stroke follows the interhemispheric inhibition (IHI) model, which attributes stroke-related motor dysfunction to an imbalance in cortical excitability between the two hemispheres of the brain. Current clinical guidelines recommend LF-rTMS targeting the contralesional primary motor cortex (M1) to improve hand motor function in stroke patients (Level A)^3^.

However, this protocol is not universally effective, especially in patients with severe motor impairments.^5^ Emerging evidence suggests that in patients with severe motor dysfunction, the contralesional motor cortex may compensate for the paralysis of the affected limb. A study applying HF-rTMS to the contralesional M1 in patients with severe hemiplegia demonstrated improved motor recovery.^6^ This finding inspired the bimodal balance recovery model, which links interhemispheric balance to lesion severity and introduces the concept of structural integrity, which refers to the preserved state of neural pathways after stroke.^7^ However, the threshold for defining structural integrity remains unclear, limiting the widespread clinical adoption of rTMS. Therefore, identifying predictors of positive rTMS response is crucial.

Brain function assessment is a potentially effective indicator for objectively predicting the efficacy of rTMS in improving stroke outcomes. For example, LF-rTMS targeting the contralesional M1 has proven more effective than HF-rTMS in stroke patients with high corticospinal tract (CST) integrity,^8^ suggesting the critical role of CST integrity in predicting post-stroke motor recovery. However, CST evaluation using diffusion tensor imaging (DTI) and other noninvasive functional imaging techniques, including functional magnetic resonance imaging (fMRI), positron emission tomography, and electroencephalography, have significant limitations, including the requirement for patient immobility, high cost, or poor spatial resolution.^9^ Functional near-infrared spectroscopy (fNIRS) has recently garnered increasing attention in rehabilitation research. As a noninvasive optical technique, fNIRS measures changes in oxygenated hemoglobin (HbO) and deoxygenated hemoglobin (HbR) concentrations in the cerebral cortex, reflecting brain activity.^10^ This method enables the construction of brain networks and monitoring complex motor learning processes without being affected by movement or posture, making it suitable for patients with stroke.^11^ Hiroaki et al. reported that patients with higher contralesional cortical HbO levels exhibited better motor recovery following LF-rTMS treatment after stroke.^12^ The authors suggested that assessing cortical lateralization using fNIRS before LF-rTMS treatment could predict therapeutic outcomes. However, no fNIRS-based predictive models have yet been developed to evaluate the effects of LF-rTMS on upper limb motor function recovery in stroke patients.

In this study, the key factors influencing the efficacy of LF-rTMS for upper limb motor recovery in stroke patients were investigated using fNIRS data. A predictive model was developed and validated to personalize rTMS treatment strategies, improve rehabilitation outcomes, and reduce healthcare cost for stroke-related upper limb motor dysfunction.

## Methods

### Participants

This prospective study comprised model development, internal validation, and external validation. Data for model development were collected from stroke patients admitted to the Department of Rehabilitation Medicine at the First Affiliated Hospital of Air Force Medical University between April 2021 and March 2023. The internal validation cohort included stroke patients admitted to the same department between July 2023 and March 2024, and the external validation cohort comprised stroke patients admitted to Xianyang Hospital of Yan’an University between June and October 2024 (Figure 1). The inclusion criteria were as follows: (1) first-time unilateral ischemic or hemorrhagic stroke confirmed via MRI or computed tomography; (2) age between 18 and 79 years; (3) within six months post-stroke; (4) stable vital signs and sufficient consciousness to participate in assessments; (5) upper extremity Fugl-Meyer assessment (UEFM) <58 for the affected arm;^13^ (6) mini-mental state examination (MMSE) score >22.^4^ The exclusion criteria included the following: (1) history of epilepsy; (2) contraindications to rTMS (for example, metal implants, pacemakers, or skull defects);^6^ (3) upper limb dysfunction caused by other neurological diseases; (4) currently taking antidepressant medication; (5) previously received rTMS treatment; (6) refusal to participate.

**Figure 1.**
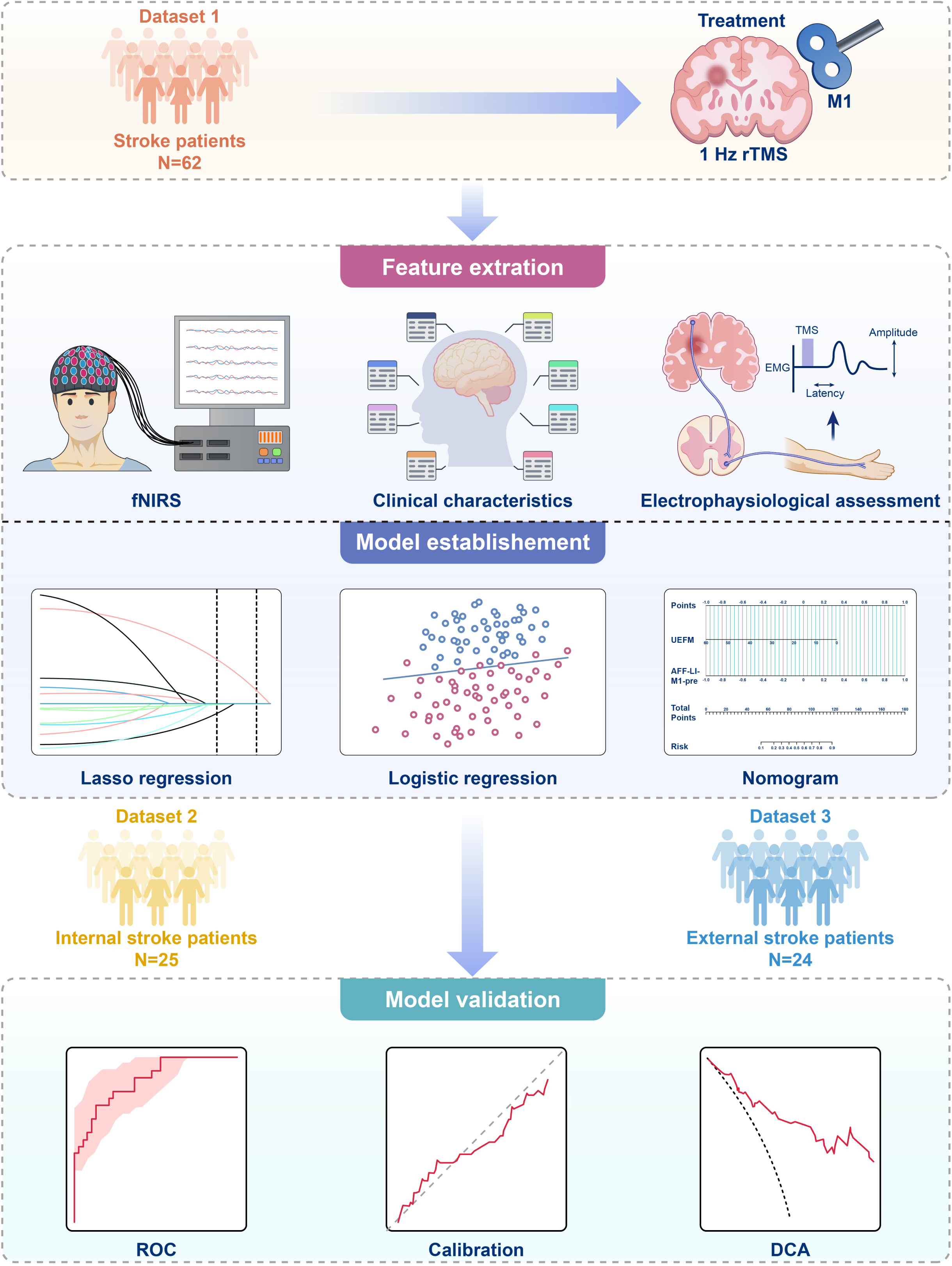
Flowchart. Abbreviations: rTMS, repetitive transcranial magnetic stimulation; M1, primary motor cortex; fNIRS: functional near-infrared spectroscopy; ROC, receiver operating characteristic; DCA, decision curve analysis; Lasso, least absolute shrinkage and selection operator.

This study represents a secondary analysis of data derived from a randomized controlled trial. The research protocol was approved by the Institutional Review Board of Xijing Hospital (approval number: KY20162079) and prospectively registered with the Chinese Clinical Trial Registry (registration number: ChiCTR-INR-17012880) prior to participant enrollment.

### Sample Size Calculation

The sample size was determined using the events-per-variable (EPV) principle, which requires at least ten outcome events per predictor in the model.^14^ Based on preliminary findings, an estimated 50% response rate to LF-rTMS treatment and three predictors for multivariate logistic regression indicated a need for 60 participants in the training cohort. According to a 7:3 ratio between the training and validation cohorts, the validation cohort required 25 participants.

### Data Collection and Assessment

Baseline data were collected before treatment, including age, gender, days post-stroke, stroke type (hemorrhagic or ischemic), brain lesion side (left or right), and lesion location (cortical, subcortical, or both). The patients were assessed by therapists with over five years of experience using the UEFM, the Wolf motor function test (WMFT), and the modified Barthel index (MBI) before and after a two-week rTMS intervention.

### Upper Extremity Fugl-Meyer Assessment (UEFM)

UEFM is a reliable and valid scale for evaluating upper limb motor impairment following stroke.^13^ It assesses motor recovery by measuring voluntary movement, coordination, muscle strength, and control, with scores ranging from 0 to 66. Higher scores indicate better motor function.

### Wolf Motor Function Test (WMFT)

WMFT is used to evaluate upper limb motor performance in daily activities using 15 standardized tasks, including grasping, moving, and fine motor movements.^13^ Tasks are scored based on time, accuracy, and ability, with a maximum total score of 75.

### Modified Barthel Index (MBI)

MBI is used to assess functional independence in activities of daily living, including eating, dressing, bathing, transferring, toileting, and walking. The scores range from 0 to 100, with higher scores indicating greater independence.^15^

The primary outcome was improvement in UEFM scores, with responders defined as those achieving a minimum clinically important difference (MCID) of ≥4.25 points.^16,17^ The secondary outcomes included changes in WMFT and MBI scores.

### Treatment

Patients underwent LF-rTMS treatment for 14 consecutive days using a magnetic stimulator (figure-eight-shaped coil, manufactured by Yiruide, Wuhan, China) tangentially positioned on the patient’s skull surface. An electromyography device was connected to the stimulator to record motor evoked potential (MEP) signals. The optimal rTMS stimulation site was defined as the location on the unaffected hemisphere that elicited the largest MEP response in the abductor pollicis brevis (APB) muscle of the unaffected upper limb. Electrophysiological assessments were performed on the APB muscle of the affected hand to evaluate MEP of the affected brain. Patients were classified as MEP-present or MEP-absent based on the presence or absence of elicited MEPs. The resting motor threshold (RMT) was determined as the minimum stimulation intensity that produced an MEP amplitude of ≥50μV in at least 5 out of 10 consecutive stimuli, delivered with an interval of >5s.^1^ The rTMS stimulation parameters were as follows: 1Hz frequency, 50s of stimulation followed by a 10s interval, targeting the hand area of the M1 region on the contralesional hemisphere at 90% RMT intensity, delivering 1000 pulses per session once daily for 14d. rTMS is reported to improve the efficacy of conventional rehabilitation therapy when applied beforehand.^1^ Consequently, rTMS treatment was performed before conventional rehabilitation sessions in this study.

All participants received conventional rehabilitation consisting of physical and occupational therapy for approximately 120min each day. Physical therapy sessions targeted posture, balance, strength, and coordination, while occupational therapy involved functional tasks (for instance, grasping objects, using chopsticks, and brushing hair) under the guidance of experienced therapists.

### fNIRS Measurement

#### fNIRS Data Acquisition

Brain activity during the fist-clenching task was measured using the NirSmart-6000A system (Huichuang Medical Equipment, China). Changes in HbO and HbR concentrations were measured across 20 channels at a sampling rate of 11 Hz, covering the M1, primary somatosensory cortex (S1), and premotor cortex (PMC). The probe array covered the regions of interest (ROIs) in both hemispheres, which are associated with motor function and play crucial roles in sensation and motor control (Figure 2A)^1^.

**Figure 2.**
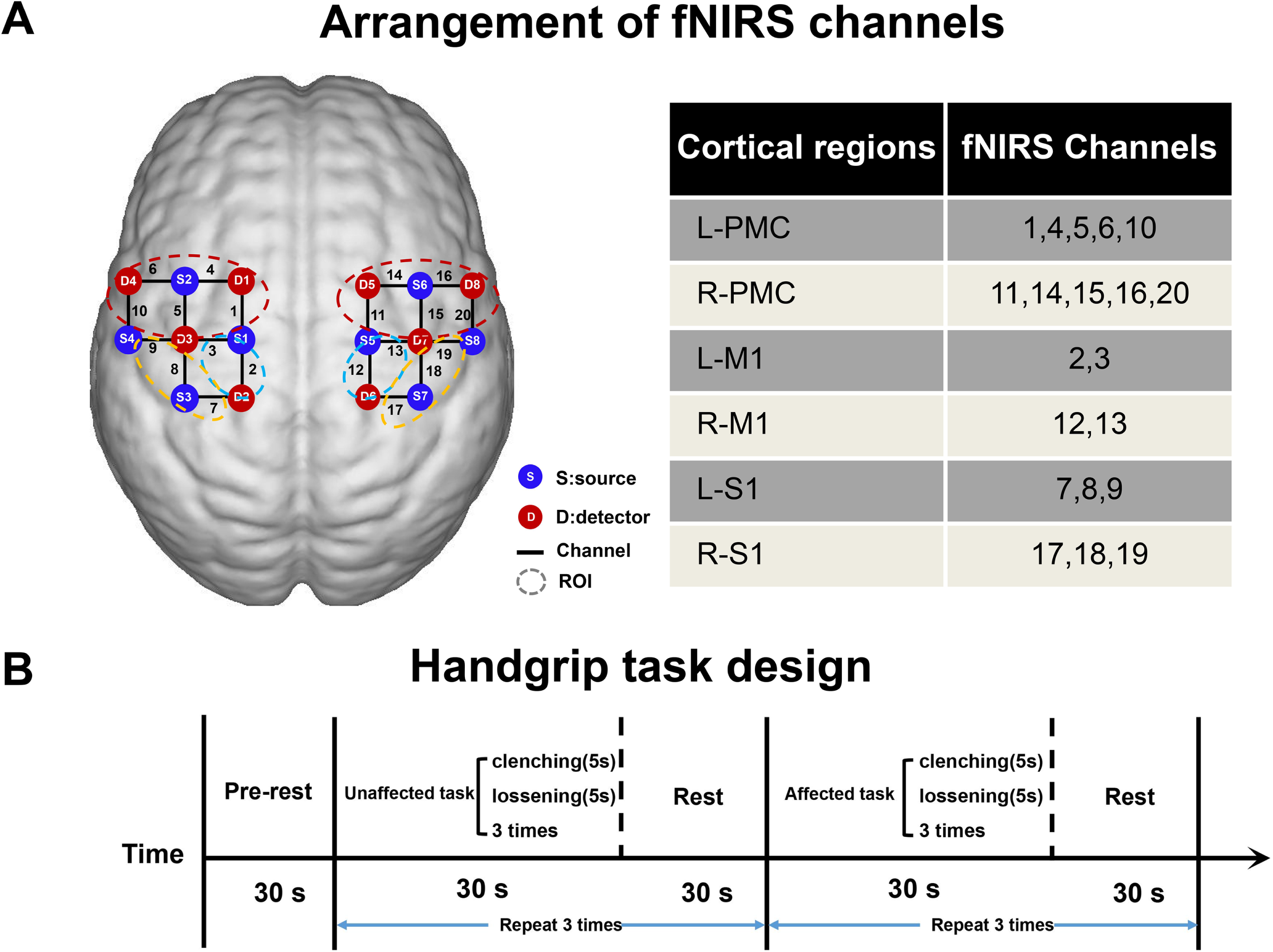
Experimental setup. (A) Layout of the fNIRS probe array. The fNIRS system includes eight detectors (red) and eight sources (blue), located according to the international 10–20 system. Dashed lines represent the ROIs, and the corresponding anatomical positions of each channel are illustrated in the table. (B) The experimental paradigm consists of three conditions: resting state, unaffected handgrip task, and affected handgrip task. Abbreviations: ROI: region of interest; fNIRS: functional near-infrared spectroscopy; L-PMC, left premotor cortex; R-PMC, right premotor cortex; L-M1, left primary motor cortex; R-M1, right primary motor cortex; L-S1, left primary somatosensory cortex; R-S1, right primary somatosensory cortex.

#### Fist-Clenching Task

The fNIRS was performed in a quiet room. Before the experiment, the patient was placed in a supine position and trained to perform the fist-clenching task. The task began with a 30s resting phase. The unaffected hand underwent three cycles of clenching the fist for 5s, followed by relaxation for 5s, and then ended with a 30s rest.^10,18^ This sequence was repeated thrice for the unaffected hand. The same procedure was applied to the affected hand (Figure 2B). If patients were unable to perform actual fist-clenching task in the affected hand, they were asked to complete motor imagery (MI) instead.^19^

#### fNIRS Data Processing

The fNIRS data were processed using the NirSpark software package (Huichuang Medical Equipment, Danyang, China). Raw light intensity data were converted to optical density data, and physiological noise (for instance, heart rate and respiration) was removed using a 0.01–0.20Hz band-pass filter. Motion artifacts were removed using a 3rd-order spline interpolation, and the signal quality was assessed using the coefficient of variation (CV) for each channel. Channels with CV >15% were excluded from further analysis to ensure data reliability. The denoised optical density data were then converted to hemoglobin concentration changes, and the average signal-to-noise ratio across all channels was calculated to confirm the quality of the processed data. HbO concentration was used for further analysis because it is a reliable and sensitive marker of motor-related brain activation.^10^ The baseline was set to [-2s, 0s]. Given that the HbO response to the fist-clenching task exhibited a 3s delay and peaked at 5s, the average HbO for each channel was calculated over the 3–7s interval following task onset.^10^ To facilitate data processing, a mirroring transformation technique was applied to the fNIRS data of stroke patients with right hemisphere lesions, converting it to represent left hemisphere lesions. The average HbO for each ROI was obtained by dividing the total HbO across all channels within the ROI by the number of channels in that ROI.^1^ To assess hemispheric activation asymmetry, the laterality index (LI) was calculated using the following formula:

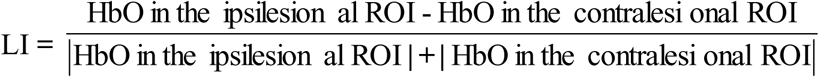

where LI >0 indicates higher activation in the ipsilesional ROI, and LI <0 indicates higher activation in the contralesional ROI.^1,12^

### Statistical Analysis

Statistical analyses were performed using the Statistical Package for the Social Sciences (version 24.0) and R (version 4.0) software. The Shapiro–Wilk test was used to assess data normality. Continuous variables are expressed as mean ± standard deviation (for normally distributed data) or median with interquartile range (for non-normally distributed data), while categorical variables are expressed as counts and percentages. Group comparisons for continuous variables were performed using independent-sample t-tests (normal data) or Mann–Whitney U tests (non-normal data), while categorical data were compared using Chi-square or Fisher’s exact tests. Paired-sample t-tests (normal data) or Wilcoxon tests (non-normal data) were used to compare pre- and post-intervention data. Pearson or Spearman correlation analyses were conducted when appropriate. To address potential model instability due to limited sample size, least absolute shrinkage and selection operator (LASSO) regression was used to preliminarily screen the factors associated with the efficacy of LF-rTMS on upper limb motor function.^20^ Multivariate logistic regression with forward stepwise selection (likelihood ratio, entry α=0.05, removal α=0.10) was used to identify the independent predictors of treatment response. A predictive model was constructed, and a nomogram was developed based on the independent predictors. The predictive performance was evaluated using the area under the receiver operating characteristic (ROC) curve (AUC), with an AUC >0.75 indicating good discriminative ability. Calibration was assessed using calibration plots, the Hosmer–Lemeshow test, and the Brier score, with lower Brier scores indicating higher prediction accuracy. The clinical utility was evaluated using decision curve analysis (DCA). *P*<0.05 was considered statistically significant.

## Results

### Participant Characteristics

A total of 98 patients were recruited into the training cohort. Based on the inclusion and exclusion criteria, 36 participants were excluded due to a history of epilepsy (n=5), contraindications to rTMS (n=25), and refusal to participate (n=6), leaving 62 patients eligible for inclusion in the study (Figure 1).

The internal validation cohort included 37 patients, of which twelve participants were excluded due to a history of epilepsy (n=1), contraindications to rTMS (n=9), and refusal to participate (n=2), leaving 25 patients for internal validation. The external validation cohort included 30 patients recruited from Yan’an University Xianyang Hospital, with six being excluded due to contraindications to rTMS (n=5) and refusal to participate (n=1), leaving 24 patients for external validation (Figure 1). Statistical analysis revealed no significant differences in demographic or clinical parameters between the training, internal, and external validation cohorts (*P*>0.05; Supplemental Table 1).

### Efficacy of LF-rTMS on Upper Limb Motor Function

After a 14-day intervention, LF-rTMS significantly increased UEFM scores (*P*<0.001; Figure 3A), WMFT scores (*P*<0.001; Supplemental Figure 1A), and MBI scores (*P*<0.001; Supplemental Figure 1B) in stroke patients. Figure 3B demonstrates the association between baseline UEFM scores and treatment-induced motor improvement (ΔUEFM=post-treatment - pre-treatment scores). The patients were classified into two groups based on whether their UEFM scores achieved the MCID after two weeks of LF-rTMS treatment: the responder (n=38) and non-responder (n=24) groups, with a response rate of 61.3%. The demographic characteristics, clinical parameters, functional assessments, and LI comparisons for each ROI between the responder and non-responder groups are detailed in Table 1. The non-responder group exhibited a significantly longer post-stroke duration than the responder group (*P*=0.049). Additionally, pre-intervention UEFM, WMFT, and MBI scores were significantly lower in the non-responder group (*P*<0.05). The proportion of patients using MI was significantly higher in the non-responder group than in the responder group (*P*=0.006). Pre-intervention, the laterality index of the primary motor cortex (LI-M1) values for the affected hand was significantly higher in the non-responder group (*P*<0.05). Using response status as the dependent variable, LASSO regression was applied to select the predictive factors from the 17 candidate variables. The results of the LASSO regression are illustrated in Supplemental Figure 2. When the model error was one standard error, three independent variables were selected: pre-intervention WMFT score (WMFT-pre), pre-intervention UEFM score (UEFM-pre), and pre-intervention laterality index for M1 for the affected hand (AFF-LI-M1-pre).

**Figure 3.**
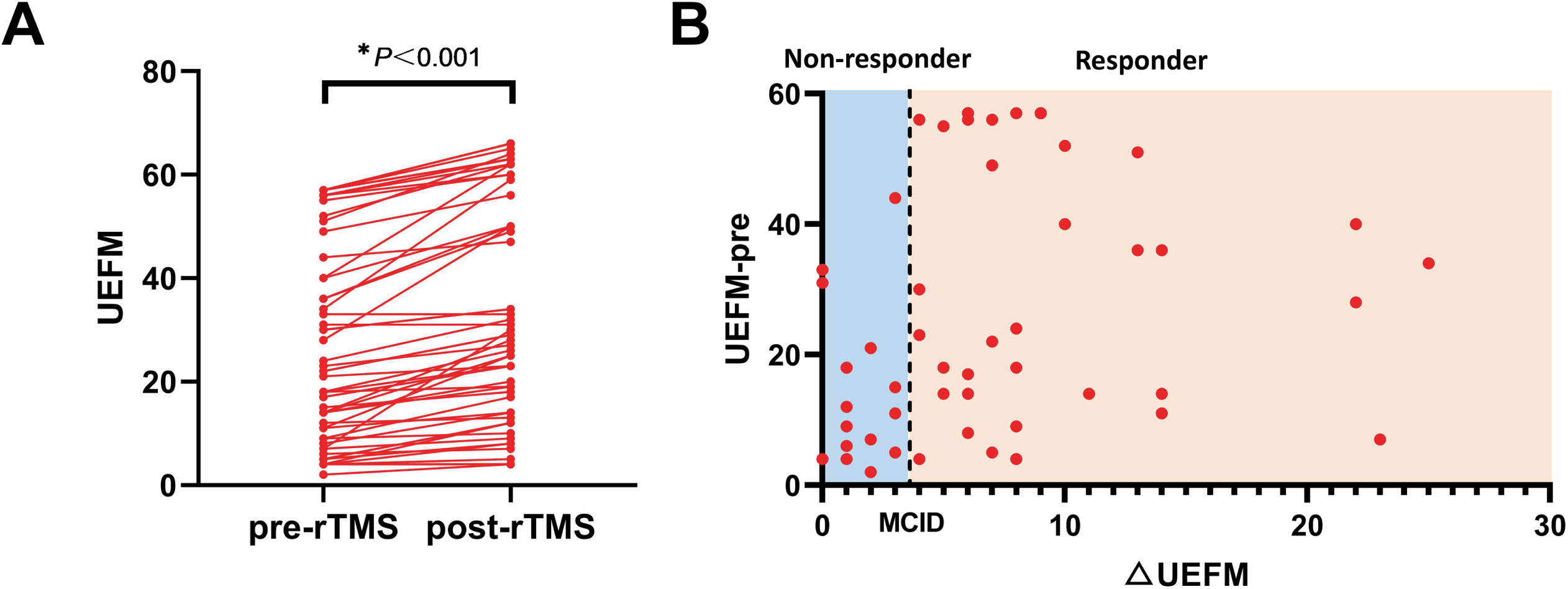
Effect of rTMS treatment on UEFM scores. (A) UEFM scores significantly improved after LF-rTMS treatment. (B) Scatter plot depicting the change in UEFM scores (△UEFM) relative to the pre-rTMS UEFM scores. The participants were divided into non-responders and responders, with the dashed line representing the MCID. Abbreviations: UEFM, Fugl-Meyer assessment upper extremity scale; rTMS, repetitive transcranial magnetic stimulation; MCID, minimum clinically important difference.

**Table 1.**
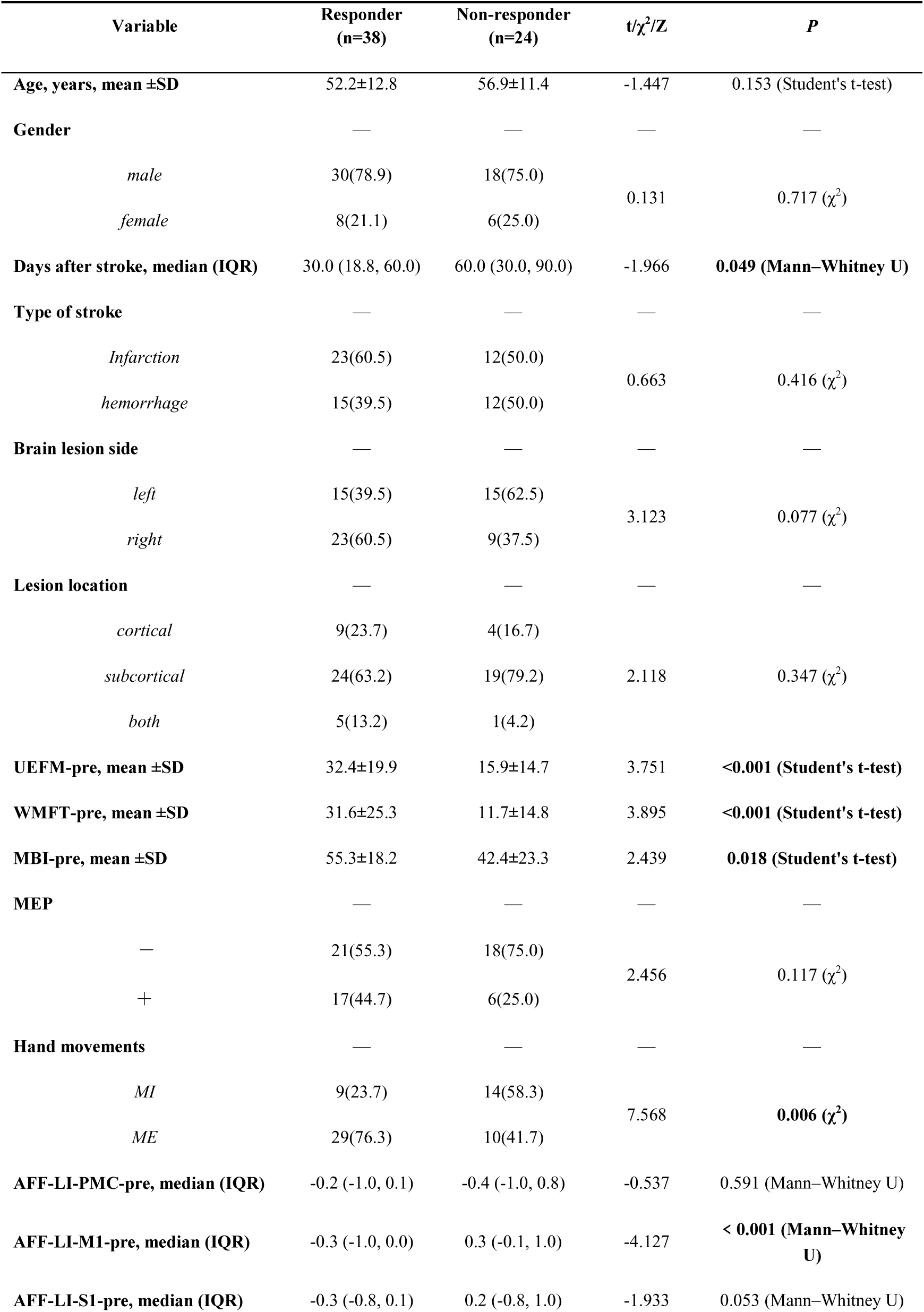
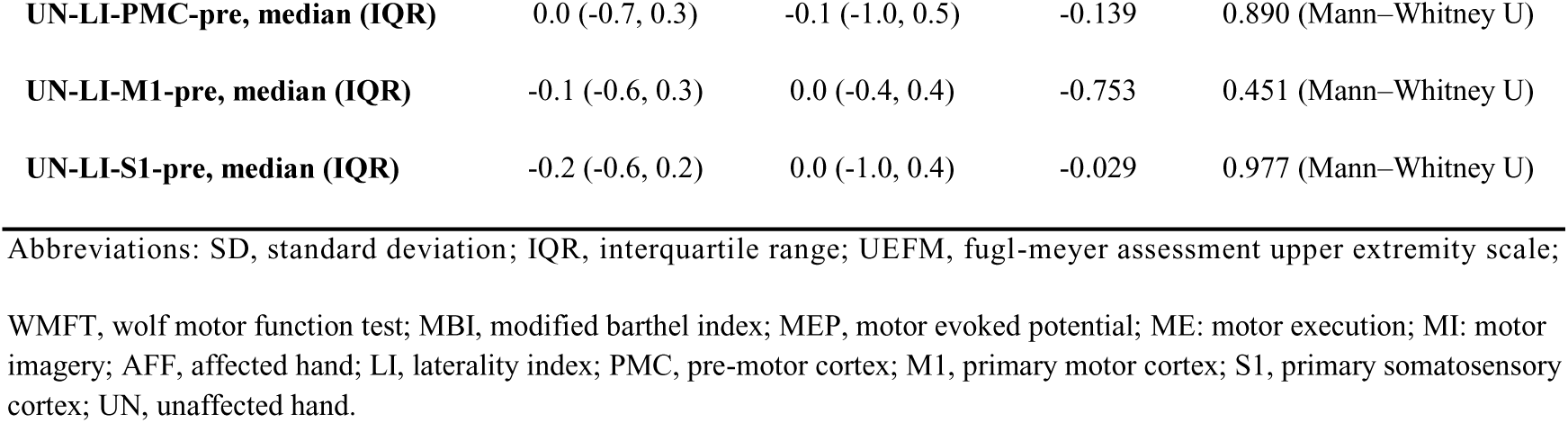
Comparison of responder and non-responder in training cohorts [n(%)].

| Variable | Responder<br>(n=38) | Non-responder<br>(n=24) | t/$\chi^2$/Z | P |
| --- | --- | --- | --- | --- |
| **Age, years, mean ±SD** | 52.2±12.8 | 56.9±11.4 | -1.447 | 0.153 (Student's t-test) |
| **Gender** | — | — | — | — |
| *male* | 30(78.9) | 18(75.0) | 0.131 | 0.717 ($\chi^2$) |
| *female* | 8(21.1) | 6(25.0) |  |  |
| **Days after stroke, median (IQR)** | 30.0 (18.8, 60.0) | 60.0 (30.0, 90.0) | -1.966 | **0.049 (Mann–Whitney U)** |
| **Type of stroke** | — | — | — | — |
| *Infarction* | 23(60.5) | 12(50.0) | 0.663 | 0.416 ($\chi^2$) |
| *hemorrhage* | 15(39.5) | 12(50.0) |  |  |
| **Brain lesion side** | — | — | — | — |
| *left* | 15(39.5) | 15(62.5) | 3.123 | 0.077 ($\chi^2$) |
| *right* | 23(60.5) | 9(37.5) |  |  |
| **Lesion location** | — | — | — | — |
| *cortical* | 9(23.7) | 4(16.7) | 2.118 | 0.347 ($\chi^2$) |
| *subcortical* | 24(63.2) | 19(79.2) |  |  |
| *both* | 5(13.2) | 1(4.2) |  |  |
| **UEFM-pre, mean ±SD** | 32.4±19.9 | 15.9±14.7 | 3.751 | **<0.001 (Student's t-test)** |
| **WMFT-pre, mean ±SD** | 31.6±25.3 | 11.7±14.8 | 3.895 | **<0.001 (Student's t-test)** |
| **MBI-pre, mean ±SD** | 55.3±18.2 | 42.4±23.3 | 2.439 | **0.018 (Student's t-test)** |
| **MEP** | — | — | — | — |
| — | 21(55.3) | 18(75.0) | 2.456 | 0.117 ($\chi^2$) |
| + | 17(44.7) | 6(25.0) |  |  |
| **Hand movements** | — | — | — | — |
| *MI* | 9(23.7) | 14(58.3) | 7.568 | **0.006 ($\chi^2$)** |
| *ME* | 29(76.3) | 10(41.7) |  |  |
| **AFF-LI-PMC-pre, median (IQR)** | -0.2 (-1.0, 0.1) | -0.4 (-1.0, 0.8) | -0.537 | 0.591 (Mann–Whitney U) |
| **AFF-LI-M1-pre, median (IQR)** | -0.3 (-1.0, 0.0) | 0.3 (-0.1, 1.0) | -4.127 | **< 0.001 (Mann–Whitney U)** |
| **AFF-LI-S1-pre, median (IQR)** | -0.3 (-0.8, 0.1) | 0.2 (-0.8, 1.0) | -1.933 | 0.053 (Mann–Whitney U) |
| **UN-LI-PMC-pre, median (IQR)** | 0.0 (-0.7, 0.3) | -0.1 (-1.0, 0.5) | -0.139 | 0.890 (Mann–Whitney U) |
| **UN-LI-M1-pre, median (IQR)** | -0.1 (-0.6, 0.3) | 0.0 (-0.4, 0.4) | -0.753 | 0.451 (Mann–Whitney U) |
| **UN-LI-S1-pre, median (IQR)** | -0.2 (-0.6, 0.2) | 0.0 (-1.0, 0.4) | -0.029 | 0.977 (Mann–Whitney U) |

Abbreviations: SD, standard deviation; IQR, interquartile range; UEFM, fugl-meyer assessment upper extremity scale;
WMFT, wolf motor function test; MBI, modified barthel index; MEP, motor evoked potential; ME: motor execution; MI: motor imagery; AFF, affected hand; LI, laterality index; PMC, pre-motor cortex; M1, primary motor cortex; S1, primary somatosensory cortex; UN, unaffected hand.

### fNIRS test results

Figure 4A depicts the activation levels of different brain regions during the fist-clenching task for the affected hand, measured before and after the intervention. No significant differences were observed in M1 activation levels on either the ipsilesional or contralesional sides before and after the intervention in the training cohort and non-responder groups (Figure 4B). However, the responder group demonstrated significantly lower post-intervention M1 activation on the contralesional side than pre-intervention levels (*P* < 0.05; Figure 4B). In the training cohort, changes in AFF-LI-M1-pre (△AFF-LI-M1-pre) were positively correlated with improvements in UEFM scores (△AFF-LI-M1-pre) (r=0.438, *P* < 0.001). Additionally, significant correlations were also observed between △AFF-LI-M1-pre and △UEFM (r=0.391, *P*=0.015) in the responder group. Conversely, no significant correlation was observed in the non-responder group (r=0.182, *P*=0.395; Figure 4C).

**Figure 4.**
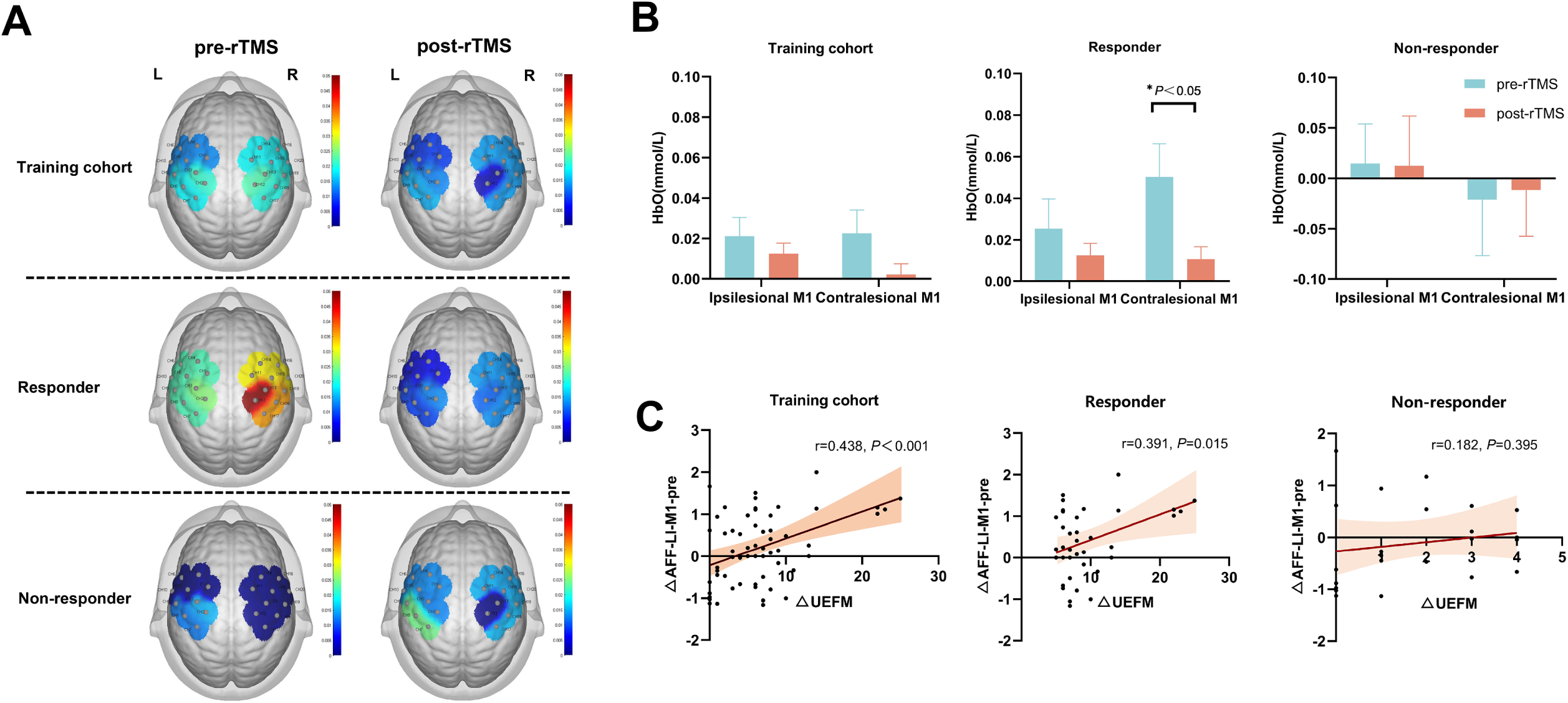
Effects of rTMS on brain activity and functional outcomes across different cohorts. (A) Mirroring transformation technique was used to convert the fNIRS data of stroke patients with right hemisphere lesions to represent left hemisphere lesions. Brain activation maps illustrating changes before and after rTMS intervention in the training cohort, responder, and non-responder groups. The maps represent the distribution of activation in the M1 regions for both the ipsilesional and contralesional hemispheres, with color scales indicating the magnitude of the change. (B) Changes in HbO levels in the training cohort, responder, and non-responder groups before and after rTMS intervention for the ipsilesional and contralesional hemispheres. (C) Correlation between △AFF-LI-M1-pre and △UEFM in the training cohort, responder, and non-responder groups. Abbreviations: rTMS, repetitive transcranial magnetic stimulation; HbO, oxyhemoglobin; L, left; R, right; M1, primary motor cortex; AFF-LI-M1-pre, pre-intervention laterality index for M1 for the affected hand; UEFM, Fugl-Meyer assessment upper extremity scale.

### Development of a nomogram

Multivariate logistic regression revealed that the UEFM-pre score (*P*=0.015, odds ratio [OR]=1.048) and AFF-LI-M1-pre (*P*=0.001, OR=0.117) were independent predictors of response to LF-rTMS (Table 2). A predictive nomogram model was developed to visualize the efficacy of LF-rTMS (Figure 5).

**Figure 5.**
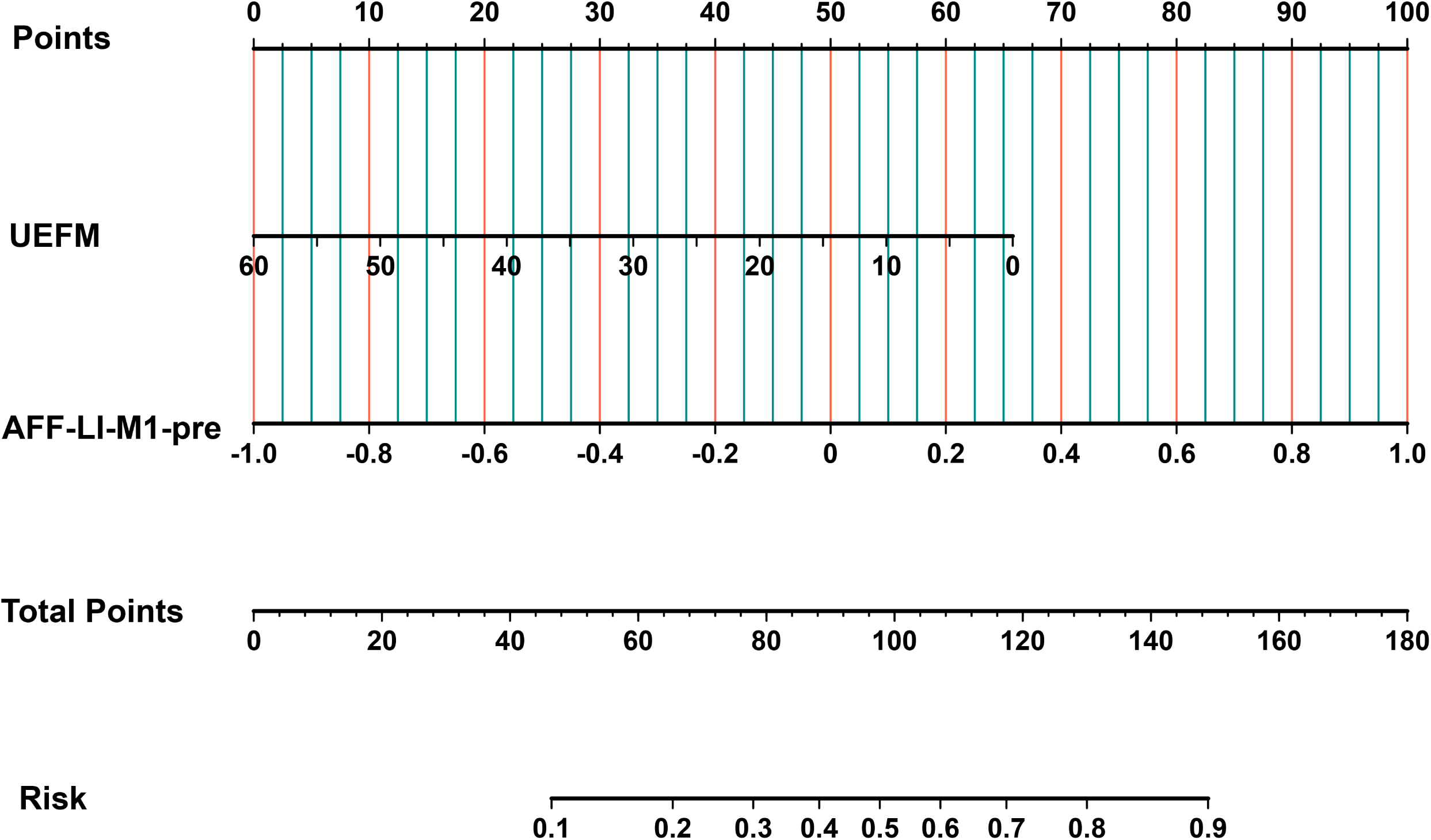
The developed nomogram. Abbreviations: UEFM, Fugl-Meyer assessment upper extremity scale; AFF-LI-M1-pre, pre-intervention laterality index for M1 for the affected hand.

**Table 2.** Multivariate logistic regression analysis of responders and non-responders in training cohorts.

| Variable | $\beta$ | wald | *P* | Odds ratio (95% CI) |
| --- | --- | --- | --- | --- |
| **UEFM-pre** | 0.047 | 5.936 | 0.015 | 1.048 (1.009 to 1.089) |
| **AFF-LI-M1-pre** | -2.145 | 11.162 | 0.001 | 0.117 (0.033 to 0.412) |

Abbreviations: UEFM, fugl-meyer assessment upper extremity scale; AFF, affected hand; LI, laterality index; M1, primary motor cortex; CI: confidence interval.

### Validation of nomogram

The ROC curve for the training cohort demonstrated good discriminatory ability, with an AUC of 0.861 (95% confidence interval [CI]: 0.770–0.952; Figure 6A). The optimal cut-off values were 34 points for the UEFM score and 0.057 for the AFF-LI-M1-pre score. The calibration curve demonstrated a strong agreement between the predicted and observed probabilities, with no significant deviation in the Hosmer–Lemeshow test (*P*=0.801; Figure 6B). The Brier score was 0.148, indicating an excellent calibration. DCA demonstrated a threshold probability range of 0.02 to 0.94, with net benefits between 7% and 38%, highlighting the nomogram’s clinical applicability (Figure 6C).

**Figure 6.**
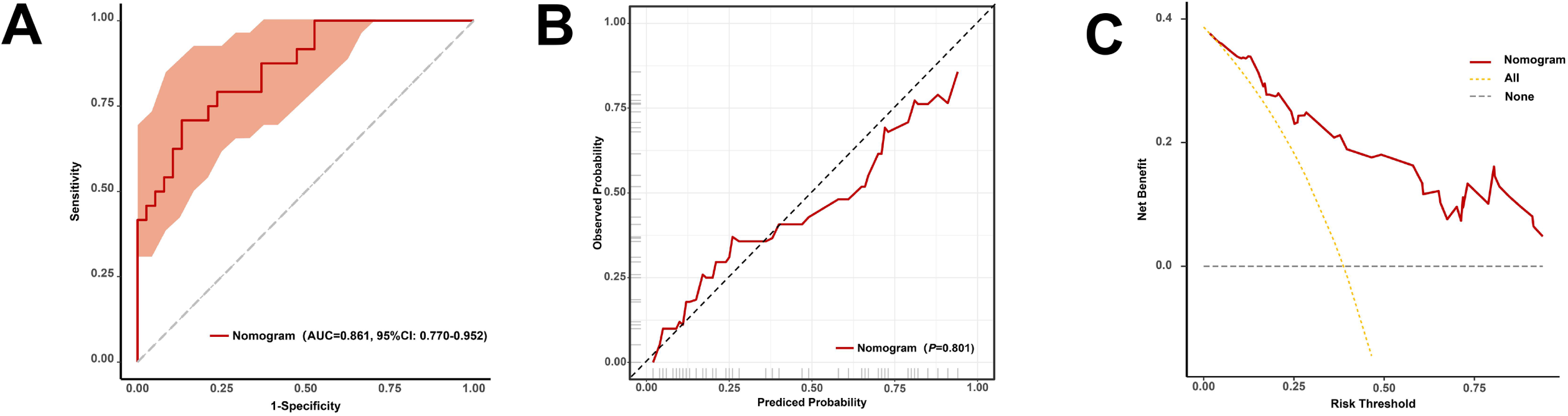
Evaluation of the nomogram prediction in the training cohort. (A) ROC curve for the nomogram, with the shaded area representing the confidence interval. (B) Calibration curve of the nomogram. (C) DCA of the nomogram. Abbreviations: ROC, receiver operating characteristic; AUC, area under the ROC curve; DCA, decision curve analysis.

The ROC curve for the internal validation cohort demonstrated an AUC of 0.853 (95% CI: 0.703–1.000), while the external validation cohort exhibited an AUC of 0.828 (95% CI: 0.645–1.000), indicating good discriminatory performance in both cohorts (Supplemental Figure 3A and D). Moreover, the calibration curves for the internal and external validation cohorts demonstrated consistency, with no significant deviation in the Hosmer–Lemeshow test (*P*=0.308 and 0.112, respectively; Supplemental Figure 3B and E). The Brier scores for the internal and external validation cohorts were 0.171 and 0.153, respectively, further supporting the excellent calibration. DCA analysis revealed threshold probabilities of 0–0.90 for the internal cohort (net benefit: 0%–40%) and 0.01–0.99 for the external cohort (net benefit: 0%–90%), indicating strong clinical applicability (Supplemental Figure 3C and F).

### Subgroup Analysis of Motor Imagery and Motor Execution Patients

A subgroup analysis was conducted to compare MI patients with motor execution (ME) patients in terms of demographics, clinical parameters, functional assessments, and LI values for each ROI (Supplemental Table 2). Pre-intervention UEFM, WMFT, and MBI scores were significantly lower in MI patients than in ME patients (*P*<0.05). Furthermore, the responder rate was significantly higher in the ME group than in the MI group (*P*=0.006). No other significant differences were observed.

ROC curve analysis indicated good discriminatory performance for independent predictors in both groups, with an AUC of 0.857 (95% CI: 0.699–1.000) for the MI group and 0.821 (95% CI: 0.679–0.963) for the ME group (Supplemental Figure 4).

## Discussion

In this study, fNIRS was used to identify cortical hemodynamic biomarkers associated with the differential responses of upper limb motor function recovery to LF-rTMS in patients with stroke. The results demonstrate that LF-rTMS effectively improved motor function and ADLs in patients with stroke. The UEFM-pre score was identified as an independent predictor of treatment efficacy; patients with better initial motor function (UEFM score ≥34 points) were more likely to benefit from LF-rTMS. Additionally, AFF-LI-M1-pre, the asymmetry of bilateral M1 activation for the affected hand measured by fNIRS, was identified as a critical predictor. Patients with lower AFF-LI-M1-pre values (<0.057) were more likely to respond to the treatment. The results of the AFF-LI-M1-pre were based on fNIRS, which is more objective than other evaluative indicators. Both internal and external validations demonstrated the stability and generalizability of the predictors. The responders exhibited decreased activation in the contralesional M1 post-intervention, indicating that LF-rTMS facilitates cortical reorganization.

The recovery of upper limb motor function in patients with stroke presents numerous challenges, particularly for those who fail to achieve full functional recovery. Developing a predictive model based on individualized patient characteristics could assist clinicians in making precise therapeutic decisions tailored to each patient’s condition. Several predictive models for post-stroke upper limb motor recovery have been proposed, with the Predict Recovery Potential (PREP) algorithm being one of the most well-established.^21,22^ The PREP algorithm primarily relies on three indicators: upper limb neurological impairment scores, the presence or absence of MEPs, and CST integrity. In clinical practice, the PREP algorithm has proven useful in guiding rehabilitation strategies, enabling physicians to develop personalized rehabilitation plans, thereby improving recovery efficiency and patients’ quality of life. However, the PREP algorithm may not be directly applicable for predicting responsiveness to rTMS. Furthermore, these algorithms typically depend on advanced equipment, such as MRI, which is not widely available in community or primary care hospitals. Therefore, the effective evaluation and prediction of the therapeutic efficacy of rTMS in resource-limited settings remains an urgent issue. In this study, the factors influencing the effects of LF-rTMS applied to the non-affected hemisphere on post-stroke upper limb motor recovery were investigated. Based on these findings, a predictive model was developed incorporating the UEFM score and LI-M1 using portable fNIRS technology, which can be implemented in community and primary care settings. Internal and external validations demonstrated the stability and generalizability of the model. These models hold significant clinical value for devising personalized rTMS treatment strategies.

The UEFM score, a standardized assessment tool for post-stroke upper limb motor function, is widely used in clinical practice and research. In this study, the pre-intervention UEFM score was identified as an independent predictor of the efficacy of LF-rTMS, with a cut-off value of 34 points. This cut-off value can serve as a critical benchmark for clinical decision-making and patient stratification. These results indicated that patients with better initial motor function (UEFM score ≥34 points) were more likely to benefit from LF-rTMS, consistent with previous studies.^23^ A systematic review indicated that the IHI theory, which posits the hypothesized mechanism for LF-rTMS stimulating the unaffected hemisphere, is contingent upon baseline motor function.^24^ Kakuda et al. investigated the potential association between the efficacy of LF-rTMS combined with occupational therapy for upper limb hemiplegia in post-stroke patients and the severity of initial hemiplegia.^23^ The study included 52 patients with stroke who were divided into three groups based on the Brunnstrom stages. All participants underwent 22 sessions of LF-rTMS treatment and 120 min of intensive occupational therapy over 15d. The results demonstrated that patients in higher Brunnstrom stages improved significantly than those in lower stages, implying that the combination of LF-rTMS and intensive occupational therapy improved upper limb motor function, with treatment efficacy influenced by the initial severity of hemiplegia. These findings highlight the significant impact of baseline motor function on the therapeutic outcomes. In the rehabilitation of stroke patients, the initial motor function level may determine the speed and magnitude of the treatment response. Patients with poorer initial function may require a longer rehabilitation period or adjustments to the treatment protocol to achieve the desired therapeutic effects. Therefore, early evaluation of a patient’s motor function status can help clinicians develop individualized treatment plans, thereby improving the precision and efficacy of rehabilitation strategies.

fNIRS can objectively predict the efficacy of LF-rTMS by assessing brain hemodynamics and hemispheric asymmetry in patients with stroke. Takeda et al. investigated cortical activation during motor tasks in five patients with cerebral infarction and five healthy controls using fNIRS. They discovered that the lateralization of primary sensorimotor cortex activation was closely associated with hand motor function in the patient group.^25^ We investigated the role of AFF-LI-M1-pre in LF-rTMS therapy and discovered that a lower AFF-LI-M1-pre value (<0.057) was a strong predictor of response to LF-rTMS. This cut-off value can serve as a critical benchmark for clinical decision-making and patient stratification. Additionally, this cut-off value can be used to identify patients who may benefit from rTMS to rebalance cortical activity. Enhanced lateralization, characterized by increased activation in the non-lesioned hemisphere than the lesioned hemisphere, is considered a marker of cortical functional asymmetry, commonly observed in stroke patients.^12^ Previous studies have demonstrated that MEP can be used to assess the lateralization of motor control in the cerebral cortex.^26^ MEP primarily focuses on assessing the CST, whereas fNIRS enables the simultaneous monitoring of multiple cortical regions, allowing for a more comprehensive evaluation of cortical activation lateralization by capturing the functional activity of numerous brain areas. Additionally, due to disruption of CST integrity, MEPs are absent on the affected side in up to 44.3% of stroke patients.^27^ Consequently, fNIRS is more effective than MEP in assessing cortical lateralization in patients with stroke. Tamashiro et al. conducted a 15-day LF-rTMS intervention targeting the non-lesioned M1 region in 59 stroke patients.^12^ Pre-intervention fNIRS assessments revealed that patients with higher non-lesioned cortical activation exhibited better motor recovery. The authors suggested that evaluating cortical asymmetry using fNIRS before initiating LF-rTMS could help predict the treatment response. However, their study only included stroke patients with Brunnstrom stage 3–6 for the affected upper limb, significantly limiting the generalizability of the results to larger clinical populations. Moreover, their research primarily focused on the primary sensorimotor cortex (BA4), premotor cortex, and supplementary motor area (BA6) as ROIs. Conversely, we accurately localized the M1 region and developed a predictive model based on this targeted approach. This refined localization not only improved the accuracy of our findings but also significantly improved the clinical utility and translational potential of our results. Furthermore, they used a single-center design, whereas we developed a predictive model that underwent internal and external validation, demonstrating its potential for enhanced clinical application.

Changes in AFF-LI-M1-pre were closely associated with improvements in upper limb motor function. These findings corroborate the mechanism by which LF-rTMS reduces the hyperexcitability of the non-affected M1 region, thereby decreasing its inhibitory effect on the affected M1 region and promoting functional recovery of the affected cerebral cortex. This mechanism aligns with the fundamental principles of cortical remodeling, particularly in the recovery process after stroke, where functional reorganization of the brain is often accompanied by a rebalance of bilateral homologous M1 activity. Nowak et al. conducted a study in which 15 stroke patients underwent 10 min of LF-rTMS on the non-affected M1 region, with fMRI assessments performed before and after the intervention.^28^ The fMRI results revealed that LF-rTMS applied to the non-affected M1 region reduced its hyperactivity. Therefore, dynamically monitoring changes in AFF-LI-M1-pre using fNIRS not only aids in identifying patients who are most likely to benefit from LF-rTMS but also facilitates the real-time evaluation of motor cortex reorganization during upper limb motor recovery.

In this study, no significant changes were observed in S1 or PMC regions, implying that M1 plays a central role in post-stroke motor recovery, whereas the contributions of S1 and PMC regions to LF-rTMS-induced motor improvements may be less pronounced. The lack of significant findings in S1 and PMC regions could be attributed to several factors. First, the primary objective of LF-rTMS in this study was to modulate cortical excitability in the M1 region, which is directly involved in motor execution. Although the S1 region is critical for sensory processing^29^ and the PMC for motor planning,^30^ their roles in the motor recovery process may be secondary to M1 activation. Second, the task paradigm used in this study (fist-clenching) primarily engages the M1 region, which may not sufficiently activate S1 or PMC to detect meaningful hemodynamic changes. Future studies should consider incorporating more complex motor or sensorimotor integration tasks to better capture the involvement of these regions. Despite the absence of significant findings in the S1 and PMC, our results highlight the critical role of M1 in motor recovery and the potential of fNIRS-derived biomarkers for predicting treatment response. Future research should investigate the interactions between M1, S1, and PMC regions, particularly in the context of different rTMS protocols or combined neuromodulation approaches.

For patients with severe upper limb motor impairment who are unable to perform actual fist-clenching tasks, MI (which involves imagining body movements while keeping the muscles at rest) was used as an alternative paradigm for fNIRS measurement. Moreover, our results demonstrated comparable predictive efficacy of the independent factors derived from both fNIRS and functional assessments between MI and ME conditions, suggesting that neuroimaging and functional data during MI may play a role akin to those during ME in forecasting treatment outcomes. Numerous studies have outlined the similarities between ME and MI. Kimberley et al. investigated the neural substrates of MI in patients with severe hemiplegia and their relationship with motor function recovery.^31^ They used fMRI to assess cortical activation patterns during wrist movement tasks (ME and MI) in 10 stroke patients and 10 healthy controls, focusing on activation differences in M1, S1, and SMA. The results demonstrated that even in cases of severe motor impairment, MI can activate functionally relevant neural networks, providing theoretical support for its application in stroke rehabilitation. Besides, MI is a fundamental theoretical basis for brain-computer interface (BCI) applications. MI-based BCI systems have been extensively used to assist patients with motor dysfunction due to stroke.^32^ A comparison of the therapeutic outcomes between MI and ME groups revealed that patients in the ME group exhibited a higher therapeutic response rate than those in the MI group. This further confirms the accuracy of the AFF-LI-M1-pre value and UEFM-pre score. The mean of AFF-LI-M1-pre in the MI group was 0.1 (>0.057), while in the ME group, it was −0.2 (<0.057). The subgroup analysis revealed that MI patients exhibited significantly lower baseline motor function (UEFM, WMFT, and MBI scores) than ME patients, which could explain the lower responder rate in the MI group. This demonstrates that patients with severe motor impairments may require tailored rTMS protocols or combined interventions (for example, MI-based BCI training) to improve treatment efficacy. Future studies should investigate the potential benefits of adjusting rTMS parameters (for instance, frequency and intensity) or integrating MI with other neuromodulation techniques to improve outcomes in this subgroup.

### Limitations

While our study established a validated predictive model through rigorous multicenter internal and external validation, several limitations warrant consideration. First, in accordance with current LF-rTMS treatment guidelines, we only included patients within 6 months post-stroke. The model’s applicability to chronic-stage patients requires further investigation. Second, although our sample size exceeded minimum requirements for model development, larger cohorts could further strengthen the generalizability of our findings. Third, our streamlined model intentionally incorporated only two key parameters (UEFM score and fNIRS-derived AFF-LI-M1-pre) - this strategic simplification enhances clinical feasibility while maintaining accuracy. The combination of objective neuroimaging biomarkers with standardized clinical assessments represents a particular strength of our approach. Future studies could explore integrating additional modalities (e.g., fMRI) while considering the trade-off between complexity and real-world implementability, especially in resource-limited settings where our current model shows particular promise.

## Conclusion

This study demonstrated the efficacy of LF-rTMS in improving upper limb motor function in patients with stroke and identified key predictors of treatment response, including UEFM-pre scores and AFF-LI-M1-pre values. Both internal and external validations demonstrated the stability and generalizability of the predictors. These findings provide a scientific basis for the individualized application of rTMS and emphasize the role of fNIRS in guiding treatment optimization.

## Data Availability

Data is available upon request to corresponding author.

## Acknowledgments

The authors express sincere appreciation to the staff in the Scientific Research Department of Danyang Huichuang Medical Co., Ltd. for their invaluable technological and academic support throughout this study. We also extend our gratitude to Home for Researchers (www.home-for-researchers.com) for their assistance with language editing.

---

**Supplemental Figure 1.** Effect of LF-rTMS on WMFT and MBI scores before and after rTMS. (A) Change in WMFT score. (B) Changes in MBI scores. Abbreviations: LF, low-frequency; rTMS, repetitive transcranial magnetic stimulation; WMFT, wolf motor function test; MBI, modified Barthel index.

**Supplemental Figure 2.** Feature selection using the LASSO binary logistic regression model. (A) Log (Lambda) values of the 17 features in the LASSO model. A coefficient profile plot was generated against the log (lambda) sequence. (B) Parameter selection in the LASSO model using ten-fold cross-validation via the minimum criterion. The partial likelihood deviation (binomial deviation) and logarithmic (lambda) curves were plotted. Using the minimum standard and 1 standard error (1-SE) of the minimum standard, a vertical dashed line was drawn at the optimal value. The optimal lambda produced three nonzero coefficients. Abbreviations: LASSO, least absolute shrinkage and selection operator; SE, standard error.

**Supplemental Figure 3.** Evaluation of the nomogram predictions during internal and external validation. (A) The ROC curve for the internal validation nomogram, with the shaded area representing the confidence interval. (B) Calibration curve of the nomogram in the internal validation. (C) DCA of the nomogram in the internal validation. (D) ROC curve for the nomogram in the external validation, with the shaded area representing the confidence interval. (E) Calibration curve of the nomogram in the external validation. (F) DCA of the nomogram in the external validation. Abbreviations: ROC, receiver operating characteristic; AUC, area under the ROC curve; DCA, decision curve analysis.

**Supplemental Figure 4.** ROC curve comparison of the performance of MI and ME. The shaded areas represent the corresponding 95% confidence intervals. Abbreviations: ROC, receiver operating characteristic; AUC, the area under the ROC curve; MI, motor imagery; ME, motor execution.

## Nonstandard Abbreviations and Acronyms

AFF-LI-M1-pre: pre-intervention LI for M1 for the affected hand
APB: abductor pollicis brevis
CST: corticospinal tract
DCA: decision curve analysis
DTI: diffusion tensor imaging
EPV: events per variable
fMRI: functional magnetic resonance imaging
fNIRS: functional near-infrared spectroscopy
HbO: oxygenated hemoglobin
HbR: deoxygenated hemoglobin
LASSO: least absolute shrinkage and selection operator
LI: laterality index
M1: primary motor cortex
MBI: modified Barthel index
MCID: minimum clinically important difference
ME: motor execution
MEP: motor evoked potential
MI: motor imagery
OR: odds ratio
PMC: premotor cortex
PREP: Predict Recovery Potential
RMT: resting motor threshold
ROC: receiver operating characteristic
ROI: region of interest
rTMS: repetitive transcranial magnetic stimulation
S1: primary somatosensory cortex
UEFM: upper extremity Fugl-Meyer assessment
WMFT: Wolf motor function test

